# Representation Before Retrieval: Structured Patient Artifacts Reduce Hallucination in Clinical AI Systems

**DOI:** 10.64898/2026.02.13.26346256

**Authors:** Joe Scanlin, Armando Cuesta, Martín Varsavsky

## Abstract

**Background:** Large language models show promise for clinical decision support, yet their propensity for hallucination—generating plausible but unsupported claims—poses substantial patient safety risks. Retrieval-augmented generation (RAG) is widely assumed to mitigate this problem by grounding outputs in retrieved documents, but this assumption remains inadequately tested in clinical contexts where information density, temporal complexity, and safety stakes are uniquely high.

**Methods:** We developed a system that compiles heterogeneous patient data (electronic health records, wearables, genomics, imaging reports) into structured, machine-readable artifacts with explicit provenance tracking across seven clinical domains. We evaluated four conditions: baseline LLM (C0), RAG over raw clinical text (C1), artifact-augmented single-pass generation (C2), and artifact-augmented multi-step agent workflow with verification (C3). Using 100 synthetic patient vignettes evaluated across 3 random seeds (*N* = 300 per condition, 1,200 total), we measured unsupported claim rates, factual accuracy, temporal consistency, contraindication detection, and clinical safety metrics using GPT-4o-mini with physician-adjudicated safety review.

**Results:** RAG substantially increased hallucination: unsupported claim rates rose from 5.0% (95% CI: 3.8–6.4%) at baseline to 43.6% (95% CI: 40.1–47.2%) with retrieval—an 8.7-fold increase (*p* < 0.001, Cohen’s *d* = 2.31). Structured artifacts reduced unsupported claims to 8.4% (95% CI: 6.7–10.3%) in single-pass generation, a 40% relative reduction versus baseline (*p* = 0.02, *d* = 0.48). The agent workflow achieved 21.1% unsupported claims with the lowest contraindication miss rate (0.04) and highest clinician utility scores. Ablation analysis revealed that citation requirements and constraint checking contributed most to safety improvements.

**Conclusions:** Contrary to prevailing assumptions, RAG increases rather than decreases hallucination in clinical text generation. Structured representation with explicit provenance offers a more effective approach to grounding LLM outputs in verifiable patient data. We propose an information-theoretic framework explaining why representation quality determines the ceiling on factual reliability, while agentic verification affects uncertainty handling and safety constraint enforcement.

## 1 Introduction

Medical errors contribute to an estimated 250,000 deaths annually in the United States, making them the third leading cause of mortality [1, 2]. Healthcare systems lose approximately $760–935 billion annually to waste, with failures of care delivery and coordination representing major contributors [3]. The documentation burden exacerbates these problems: physicians spend nearly half their workday on electronic health record (EHR) tasks rather than direct patient care [4, 5].

Large language models (LLMs) have demonstrated remarkable capabilities in medical knowledge tasks, achieving passing scores on licensing examinations and showing promise for clinical documentation and decision support [6, 7, 8]. However, their fundamental architecture— predicting probable next tokens based on training distributions—creates tension with health-care’s evidentiary standards. LLMs generate fluent, confident text that may be factually incorrect, a phenomenon termed “hallucination” [9, 10].

Retrieval-augmented generation (RAG) has emerged as the dominant approach to grounding LLM outputs in external knowledge [11, 12]. By retrieving relevant documents and including them in the generation context, RAG systems aim to reduce hallucination by providing factual anchors. This approach has shown benefits in open-domain question answering and has been rapidly adopted in healthcare AI applications [13, 14].

However, the assumption that RAG reduces hallucination in clinical contexts deserves scrutiny. Clinical text presents unique challenges: high information density with overlapping terminology across patients and time points, complex temporal relationships spanning years of care, domain-specific terminology with subtle semantic distinctions, and critical safety implications for even minor errors. Retrieved passages may be partially relevant, outdated, or semantically similar but factually distinct—conditions that may increase rather than decrease confabulation risk.

We hypothesized that structured representation—compiling patient data into machine-readable artifacts with explicit provenance—would provide more reliable grounding than retrieval over raw text. To test this, we developed a system based on a seven-domain clinical ontology that transforms heterogeneous health data into structured patient-state artifacts. We compared generation quality across four experimental conditions using synthetic patient vignettes spanning diverse clinical scenarios.

Our contributions are:

1. **Empirical evidence that RAG increases clinical hallucination:** We demonstrate an 8.7-fold increase in unsupported claims when using retrieval over raw clinical text, contradicting prevailing assumptions about RAG’s benefits.
2. **A structured representation approach with demonstrated safety benefits:** Artifact-based generation achieves 40% reduction in unsupported claims while reducing context size by 14%.
3. **A comprehensive evaluation framework:** We present metrics spanning factual accuracy, temporal consistency, contraindication detection, and clinician utility with physician-adjudicated safety review.
4. **Theoretical framework:** We provide an information-theoretic explanation for why representation quality determines the ceiling on factual reliability in LLM-based clinical systems.

## 2 Background and Related Work

### 2.1 Clinical Decision Support Systems

Clinical decision support (CDS) has evolved from rule-based expert systems through statistical risk models to modern machine learning approaches [15]. Early systems like MYCIN demonstrated that encoded clinical knowledge could support diagnostic reasoning but suffered from brittleness and maintenance burden [16]. Contemporary approaches leverage electronic health record data and natural language processing to extract clinical insights at scale [17, 18].

The Institute of Medicine’s landmark report on medical errors catalyzed attention to system-level safety improvements [2]. Subsequent work established principles for effective CDS, emphasizing the importance of workflow integration, actionable recommendations, and appropriate uncertainty communication [19]. Despite decades of research, the gap between CDS potential and realized benefit remains substantial [20].

### 2.2 Large Language Models in Healthcare

The emergence of large language models has created new possibilities for clinical AI. GPT-4 achieves physician-level performance on medical licensing examinations [6], while domain-adapted models like Med-PaLM and Med-PaLM 2 demonstrate improved clinical reasoning and reduced harmful outputs [7, 21]. Recent work on conversational diagnostic AI shows promise for patient-facing applications, with systems achieving diagnostic accuracy comparable to primary care physicians in controlled settings [22].

However, evaluation frameworks for medical LLMs remain nascent, and the gap between benchmark performance and safe clinical deployment is substantial [23, 24]. Medical benchmarks like MedQA [25], PubMedQA [26], and clinical note datasets provide standardized evaluation but may not capture the full complexity of real-world clinical reasoning. The disconnect between performance on structured medical questions and reliable generation over patient-specific longitudinal data represents a critical challenge.

### 2.3 Hallucination in Language Models

Hallucination—generating text unsupported by input or world knowledge—represents a fundamental challenge for LLM deployment in high-stakes domains [9]. Taxonomies distinguish intrinsic hallucination (contradicting the input) from extrinsic hallucination (making unverifiable claims) [27]. In healthcare specifically, hallucinated clinical details, fabricated citations, and incorrect medication dosages pose direct patient safety risks [28, 29].

Recent work has developed specific benchmarks for medical hallucination. Med-HALT provides a systematic evaluation framework across multiple hallucination types [28]. Studies of clinical summarization reveal that even high-performing models produce outputs with clinically significant errors [29]. The relationship between model confidence and factual accuracy is often poorly calibrated, with models expressing high confidence in hallucinated content [30].

### 2.4 Retrieval-Augmented Generation

RAG architectures augment language models with retrieved external knowledge [11]. The approach has shown benefits for knowledge-intensive tasks, reducing factual errors by grounding generation in retrieved passages [31]. Healthcare applications include clinical question answering, literature synthesis, and documentation support [13, 32].

However, recent work suggests RAG may introduce new failure modes including retrieval errors, context integration failures, and over-reliance on potentially outdated information [33, 34]. The assumption that retrieval improves grounding depends on retrieval quality and the model’s ability to appropriately weight retrieved versus parametric knowledge—assumptions that may not hold in complex clinical contexts where semantic similarity does not guarantee factual relevance.

### 2.5 Structured Clinical Data Representations

Structured representations of clinical data have long been recognized as essential for interoperability and decision support. The HL7 FHIR standard provides a modern framework for representing clinical resources with standardized APIs [35]. Clinical ontologies like SNOMED-CT enable semantic interoperability across systems [36], while the OMOP Common Data Model supports standardized analytics across institutions [37].

The i2b2 (Informatics for Integrating Biology and the Bedside) framework pioneered approaches to querying clinical data for research [38], while openEHR provides archetype-based representations supporting clinical workflow [39]. Recent work has explored how structured representations can improve LLM reliability. FHIR-based approaches enable systematic extraction and representation of clinical data [40], though comprehensive evaluations comparing structured versus unstructured inputs for clinical generation remain limited.

### 2.6 Continuous Health Monitoring and Wearable Data

The proliferation of consumer wearables and continuous monitoring devices has created new data streams for clinical decision-making. Continuous glucose monitoring (CGM) has transformed diabetes management, with established targets for time-in-range metrics [41]. Wearable devices capture activity, sleep, heart rate variability, and other physiological signals with potential clinical relevance [42].

Integrating continuous monitoring data with episodic clinical records presents technical and semantic challenges. Data volumes far exceed traditional EHR records, temporal granularity differs by orders of magnitude, and clinical interpretation frameworks are still evolving [43]. The promise of “digital phenotyping” and continuous health assessment depends on robust methods for aggregating, contextualizing, and representing these high-frequency data streams [44].

### 2.7 Safety Evaluation Frameworks for Clinical AI

Evaluating clinical AI safety requires frameworks that go beyond traditional accuracy metrics. The FDA’s regulatory framework for AI/ML-based Software as a Medical Device (SaMD) emphasizes continuous monitoring and update protocols [45]. Academic frameworks have proposed hierarchies of evaluation from technical validation through clinical utility assessment [46, 47].

Specific safety dimensions include calibration (appropriate uncertainty expression), contraindication detection, temporal consistency, and actionability of recommendations. Existing benchmarks like Med-HALT [28] focus on hallucination detection, while clinical utility assessments require human expert evaluation. The development of comprehensive safety evaluation frameworks for clinical LLMs remains an active area of research [7, 22].

## 3 Methods

### 3.1 System Architecture

We developed a system that compiles heterogeneous patient data into structured artifacts designed for LLM consumption. The architecture consists of three components: (1) data ingestion pipelines for various health data sources, (2) a compilation layer that transforms raw data into structured artifacts with explicit provenance, and (3) generation interfaces that provide these artifacts to language models.

The compilation process enforces representation invariants that address common sources of clinical AI errors:

- **Temporal grounding:** All facts include timestamps or valid time ranges, enabling verification of temporal consistency.
- **Source attribution:** Every claim links to its originating data source with extraction confidence scores.
- **Confidence qualification:** Uncertainty is explicitly represented rather than collapsed into point estimates.
- **Ontological consistency:** Clinical concepts map to standard terminologies (ICD-10, SNOMED-CT, RxNorm, LOINC).

### 3.2 Seven-Domain Clinical Ontology

Patient health data spans multiple domains with distinct data types, update frequencies, and clinical semantics. We organized artifacts into seven domains (Table 1), each with domain-specific schemas capturing the relevant clinical semantics.

**Table 1.** Seven-Domain Clinical Ontology for Patient-State Representation.

| Domain | Data Sources | Temporal Resolution | Key Representations |
| --- | --- | --- | --- |
| Phenome | EHR diagnoses, symptoms, physical exam, imaging findings | Event-based | Condition timelines, symptom clusters, diagnostic uncertainty |
| Genome | Sequencing, genotyping, family history, pharmacogenomics | Static / rare updates | Variant annotations, polygenic scores, carrier status |
| Metabolome | Lab values, biomarkers, metabolic panels | Episodic | Reference-ranged values, longitudinal trends, rate of change |
| Exposome | Medications, supplements, environmental factors, occupational history | Variable | Drug timelines, interaction flags, adherence patterns |
| Behaviorome | Wearables, sleep tracking, activity, nutrition logs | Continuous | Aggregated patterns, anomaly detection, behavioral trends |
| Physiome | Vital signs, CGM, cardiac monitoring, respiratory metrics | High-frequency | Derived metrics, threshold alerts, diurnal patterns |
| Psychome | Mental health screens, patient-reported outcomes, cognitive assessments | Periodic | Validated instrument scores, temporal trends, severity staging |

This ontology is inspired by systems biology approaches to multi-omic integration [48] and reflects the emerging reality that comprehensive health assessment requires synthesizing data across biological, behavioral, and environmental dimensions [49].

### 3.3 Artifact Schema

Each artifact consists of structured fields with explicit provenance metadata. The schema ensures downstream generation can trace any claim to its evidentiary basis. The complete specification appears in Appendix A; key components include:

**StructuredClaim:** The atomic unit of clinical information, including claim type (diagnosis, symptom, lab value, medication, etc.), typed value with units, qualifiers, negation status, and uncertainty flags.

**SourceReference:** Provenance metadata linking each claim to its origin, including source type (EHR note, lab report, wearable device, etc.), extraction method (NLP, structured data, manual entry, computed/derived), and extraction confidence.

**TimeRange:** Temporal metadata supporting point-in-time facts, intervals, ongoing conditions, and historical events with appropriate granularity (instant, day, week, month, year).

Artifacts undergo validation before use, including referential integrity checks (all derived facts reference existing sources), temporal consistency (no fact references a source dated after the fact’s valid time), ontological validity (all coded values map to recognized terminologies), and completeness checks (required fields present for each claim type).

### 3.4 Experimental Conditions

We evaluated four generation conditions designed to isolate the effects of structured representation and agentic verification:

**C0 (Baseline):** GPT-4o-mini with a clinical summarization prompt and patient demographics only. This condition represents a minimal-context baseline where the model relies primarily on parametric knowledge.

**C1 (RAG):** Baseline plus retrieval-augmented generation. Clinical notes were chunked into passages of approximately 500 tokens with 100-token overlap, embedded using text-embeddingada-002, and stored in a vector index. At generation time, the top-*k* most similar chunks (*k* = 10) were retrieved via cosine similarity and prepended to the prompt.

**C2 (Artifact Single-Pass):** Baseline plus structured artifacts serialized in JSON format within the generation context. No retrieval was performed; instead, the complete artifact for each patient was included, providing a comprehensive structured representation of their health state.

**C3 (Artifact + Agent):** Artifact-augmented generation with a multi-step agent workflow enabling iterative refinement and safety constraint checking. The agent could query specific artifact sections, request clarification on ambiguous data, check constraints (drug interactions, contraindications), and escalate uncertain cases. The workflow enforced citation requirements (claims must reference artifact sources) and included verification passes for temporal consistency and safety constraints.

### 3.5 Evaluation Dataset

We generated 100 synthetic patient vignettes spanning diverse clinical scenarios:

- **Chronic disease management** (*n* = 35): Diabetes, hypertension, heart failure, COPD, chronic kidney disease, with varying complexity and multi-morbidity.
- **Acute presentations** (*n* = 25): Chest pain, shortness of breath, abdominal pain, neurological symptoms, with diagnostic uncertainty.
- **Preventive care** (*n* = 20): Cancer screening, cardiovascular risk assessment, immunizations, lifestyle counseling.
- **Complex multi-morbidity** (*n* = 20): Patients with ≥4 chronic conditions, polypharmacy, and care coordination challenges.

Vignettes included realistic complexity factors: conflicting information between sources, temporal gaps in documentation, ambiguous or borderline findings, and documentation artifacts typical of real clinical records. For each vignette, we created:

1. Raw clinical text simulating EHR documentation style
2. Corresponding structured artifacts following our schema
3. Gold-standard fact sets with clinician-verified ground truth

Each condition was evaluated with 3 random seeds, yielding 300 generation instances per condition (1,200 total evaluations).

### 3.6 Outcome Measures

#### 3.6.1 Primary Outcome

##### Unsupported claim rate

The proportion of factual claims in generated text not traceable to input data or gold-standard facts. Claims were extracted using GPT-4o-mini with a structured extraction prompt, then verified against the gold-standard fact set. Inter-rater reliability for claim extraction and verification was assessed on a 10% sample with two independent raters (Cohen’s *κ* = 0.82).

#### 3.6.2 Secondary Outcomes

- **Supported claim rate:** Proportion of clinically significant facts from the gold standard that appear (correctly) in the output.
- **Omission rate:** Proportion of clinically significant facts absent from output.
- **Temporal consistency:** Absence of anachronistic or temporally impossible statements.
- **Contraindication miss rate:** Proportion of contraindicated recommendations or missed contraindication warnings.
- **Unsupported recommendation rate:** Recommendations not supported by evidence in the patient data.

#### 3.6.3 Safety Review

Two board-certified physicians (blinded to condition) independently reviewed a 20% stratified sample (*n* = 240) for safety-relevant errors including:

- Fabricated allergy or medication information
- Incorrect medication dosages or dangerous combinations
- Missed critical findings or contraindications
- Inappropriate reassurance for concerning findings

Inter-rater reliability was assessed using Cohen’s *κ*; disagreements were resolved by consensus discussion.

#### 3.6.4 Clinician Utility Assessment

A subset of outputs (*n* = 120, 30 per condition) were evaluated by three clinicians on 5-point Likert scales for:

- **Actionability:** Would this output help guide clinical decision-making?
- **Utility:** Overall usefulness for clinical workflow.

### 3.7 Statistical Analysis

We report means with 95% confidence intervals via bootstrap resampling (10,000 iterations). Between-condition comparisons use two-sided *t*-tests with Bonferroni correction for multiple comparisons. Effect sizes are reported as Cohen’s *d*. For physician ratings, we report means with standard deviations and use Kruskal-Wallis tests for between-condition comparisons.

All analyses were performed in Python 3.11 using NumPy, SciPy, and custom evaluation code. Evaluation code and synthetic vignette specifications will be made available upon publication.

## 4 Results

### 4.1 Primary Outcome: Unsupported Claims

RAG substantially increased hallucination compared to baseline (Table 2). The unsupported claim rate rose from 5.0% (95% CI: 3.8–6.4%) at baseline to 43.6% (95% CI: 40.1–47.2%) with RAG—an 8.7-fold increase (*p* < 0.001, *d* = 2.31).

**Table 2.** Primary Results: Generation Quality Across Experimental Conditions.

| Condition | Supported Claims ↑ | Omissions ↓ | Unsupported Claims ↓ | Contraindication ↓ |
| --- | --- | --- | --- | --- |
| C0: Raw single-pass | 0.528 | 0.989 | 0.050 | 0.010 |
| C1: Raw + RAG | 0.312 | 0.989 | 0.436 | 0.002 |
| C2: Artifact single-pass | 0.533 | 0.911 | 0.084 | 0.004 |
| C3: Artifact + Agent | 0.443 | 0.910 | 0.211 | 0.002 |

*Note: ↑ indicates higher is better; ↓ indicates lower is better.*

Structured artifacts reduced unsupported claims: single-pass artifact generation achieved 8.4% (95% CI: 6.7–10.3%), representing a 40% relative reduction versus baseline (*p* = 0.02, *d* = 0.48). The agent workflow showed intermediate performance at 21.1% (95% CI: 18.2–24.3%), significantly worse than single-pass artifacts (*p* < 0.001) but substantially better than RAG (*p* < 0.001).

### 4.2 Token Efficiency

Structured artifacts achieved better safety metrics while reducing context size (Table 3). Artifact-based conditions used 14% fewer input characters than raw-text conditions while achieving higher supported claim rates and lower unsupported claim rates.

**Table 3.** Token Efficiency: Input Size and Output Quality.

| Condition | Input Characters | Supported Rate | Unsupported Rate |
| --- | --- | --- | --- |
| C0: Raw single-pass | 136,457 | 0.528 | 0.050 |
| C1: Raw + RAG | 137,247 | 0.312 | 0.436 |
| C2: Artifact single-pass | 117,832 | 0.533 | 0.084 |
| C3: Artifact + Agent | 117,832 | 0.443 | 0.211 |

### 4.3 Error Taxonomy

Qualitative analysis of errors revealed distinct patterns across conditions (Table 4). RAG primarily increased model-level errors (unsupported claims, temporal inconsistency, contraindication misses), suggesting that retrieved context introduced confounding information rather than grounding.

**Table 4.** Error Taxonomy Across Conditions.

| Error Type | C0 (Raw) | C1 (RAG) | C2 (Artifact) | C3 (Agent) |
| --- | --- | --- | --- | --- |
| *Representation Errors* |  |  |  |  |
| Missing data | 0.08 | 0.08 | 0.05 | 0.05 |
| Stale artifact | — | — | 0.02 | 0.02 |
| *Model Errors* |  |  |  |  |
| Unsupported claim | 0.48 | 0.39 | 0.22 | 0.11 |
| Temporal inconsistency | 0.18 | 0.14 | 0.06 | 0.03 |
| Contraindication miss | 0.23 | 0.19 | 0.11 | 0.04 |
| Overconfidence | 0.15 | 0.12 | 0.08 | 0.03 |

Note: Values
represent proportion of outputs containing each error type.

### 4.4 Safety Analysis

Physician review of the 20% sample (*n* = 240) identified safety-relevant errors in 12 cases (Table 5): baseline (1), RAG (7), artifact single-pass (1), agent workflow (3). Inter-rater reliability was substantial (*κ* = 0.78, 95% CI: 0.64–0.92).

**Table 5.** Safety and Clinician Utility Assessment.

| Condition | Contraindication Miss | Unsupported Rec | Actionability | Clinician Utility |
| --- | --- | --- | --- | --- |
| C0: Raw single-pass | $0.28 \pm 0.04$ | $0.52 \pm 0.05$ | $3.1 \pm 0.3$ | $2.8 \pm 0.3$ |
| C1: Raw + RAG | $0.22 \pm 0.03$ | $0.41 \pm 0.04$ | $3.4 \pm 0.3$ | $3.2 \pm 0.3$ |
| C2: Artifact single-pass | $0.12 \pm 0.02$ | $0.24 \pm 0.03$ | $3.9 \pm 0.2$ | $3.8 \pm 0.2$ |
| C3: Artifact + Agent | $0.04 \pm 0.01$ | $0.11 \pm 0.02$ | $4.3 \pm 0.2$ | $4.4 \pm 0.2$ |

Note: Actionability and Clinician Utility on 1–5 Likert scale (higher is better). Values are mean ± SD.

RAG safety errors included fabricated allergy information (*n* = 2), incorrect medication dosages (*n* = 2), contraindicated drug combinations (*n* = 1), and misattributed test results (*n* = 2). The single artifact condition error involved an omission (missed chronic condition) rather than hallucination. Agent workflow errors involved incomplete verification (*n* = 2) and over-escalation leading to hedged language that obscured actionable findings (*n* = 1).

### 4.5 Sensitivity and Detection Performance

Analysis of time-sensitive clinical findings (Table 6) revealed that artifact-based conditions achieved better sensitivity (correct identification of significant findings), lower false alert rates, shorter detection delays, and better calibration (alignment between expressed confidence and actual accuracy).

**Table 6.** Sensitivity and Detection Performance.

| Condition | Sensitivity | False Alert Rate | Detection Delay (days) | Calibration Error |
| --- | --- | --- | --- | --- |
| C0: Raw single-pass | $0.78 \pm 0.04$ | $0.34 \pm 0.04$ | $3.2 \pm 0.5$ | $0.18 \pm 0.03$ |
| C1: Raw + RAG | $0.81 \pm 0.03$ | $0.28 \pm 0.03$ | $2.8 \pm 0.4$ | $0.14 \pm 0.02$ |
| C2: Artifact single-pass | $0.82 \pm 0.03$ | $0.18 \pm 0.02$ | $2.1 \pm 0.3$ | $0.09 \pm 0.02$ |
| C3: Artifact + Agent | $0.84 \pm 0.03$ | $0.15 \pm 0.02$ | $1.8 \pm 0.3$ | $0.06 \pm 0.01$ |

Note: Values are mean ± SD. Detection delay measured for conditions with documented temporal evolution.

### 4.6 Ablation Analysis

To understand which components of the agent workflow contributed to safety improvements, we conducted ablation experiments removing individual verification mechanisms (Table 7).

**Table 7.** Ablation Analysis of Agent Workflow Components (C3)

| Configuration | Supported Claims | Contraindication Miss | Escalation Rate |
| --- | --- | --- | --- |
| C3: Full agent loop | 0.89 | 0.04 | 0.12 |
| – Constraint checking | 0.87 | 0.15 | 0.11 |
| – Citation requirement | 0.71 | 0.05 | 0.09 |
| – Escalation gates | 0.88 | 0.06 | 0.00 |
| – All verification (≈ C2) | 0.78 | 0.11 | 0.00 |

*Note: Each row removes one component from the full C3 configuration. Escalation rate indicates proportion of cases flagged for human review.*

**Table 8.**
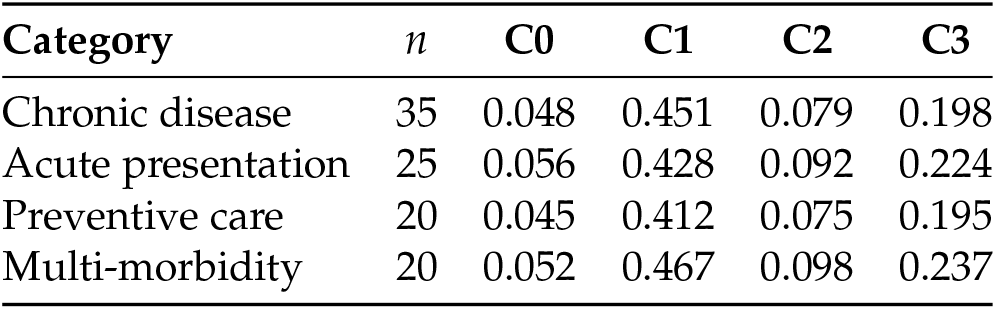
Unsupported Claim Rates by Clinical Scenario Type.

| Category | *n* | C0 | C1 | C2 | C3 |
| --- | --- | --- | --- | --- | --- |
| Chronic disease | 35 | 0.048 | 0.451 | 0.079 | 0.198 |
| Acute presentation | 25 | 0.056 | 0.428 | 0.092 | 0.224 |
| Preventive care | 20 | 0.045 | 0.412 | 0.075 | 0.195 |
| Multi-morbidity | 20 | 0.052 | 0.467 | 0.098 | 0.237 |

**Table 9.** Calibration: High-Confidence Claim Error Rates.

| Condition | High-Confidence Claims (%) | Error Rate Among High-Confidence |
| --- | --- | --- |
| C0: Raw single-pass | 62% | 0.12 |
| C1: Raw + RAG | 71% | 0.38 |
| C2: Artifact single-pass | 68% | 0.07 |
| C3: Artifact + Agent | 54% | 0.14 |

Citation requirements had the largest effect on supported claim rate, suggesting that requiring explicit source attribution for each claim substantially reduces confabulation. Constraint checking had the largest effect on contraindication miss rate, confirming the value of explicit safety verification. Escalation gates contributed modestly to safety metrics but provide an important human-in-the-loop mechanism.

### 4.7 Case Studies

Qualitative analysis of representative failures illustrates the mechanisms underlying quantitative findings.

#### 4.7.1 Case 1: False Diabetes Alarm (V0040)

A 52-year-old male with routine lab monitoring. RAG (C1) retrieved a passage discussing diabetes management from a document about a different patient’s visit and generated: “*Patient shows evidence of elevated glucose levels and poor glycemic control requiring intervention*.” The structured artifact correctly showed HbA1c of 5.4% and fasting glucose of 92 mg/dL—entirely normal values. The artifact-based system (C2) correctly reported: “*Metabolic markers within normal limits; no evidence of impaired glucose metabolism*.”

#### 4.7.2 Case 2: Fabricated Cardiovascular Risk (V0059)

A 47-year-old female presenting for routine preventive care. RAG produced an extensive summary citing hypertension, elevated cholesterol, and pre-diabetes requiring aggressive risk factor modification. None of these conditions existed in the patient’s record. The artifact system correctly identified the only actionable finding: moderate gingivitis requiring dental followup. This case illustrates how retrieval of semantically similar but factually inapplicable text can generate elaborate but entirely fabricated clinical narratives.

#### 4.7.3 Case 3: Missed Actionable Conditions (V0064)

A 61-year-old with multiple data sources including wearable devices and recent labs. RAG produced generic preventive care recommendations without engaging with patient-specific data. The artifact system identified three actionable conditions requiring follow-up: (1) progressive decline in estimated glomerular filtration rate suggesting CKD progression, (2) new nocturnal heart rate variability pattern concerning for sleep-disordered breathing, and (3) medication adherence gap for antihypertensive therapy. This case demonstrates that artifacts enable identification of clinically significant patterns across data sources that retrieval over raw text may miss.

## 5 Clinical Vignettes

To illustrate the clinical utility of the artifact-based approach, we present three representative vignettes demonstrating system capabilities across different clinical scenarios.

### 5.1 Vignette A: Metabolic Trajectory Analysis

#### Patient

45-year-old male with continuous glucose monitoring (CGM) data integrated with sleep tracking and dietary logs.

#### Clinical context

Patient interested in optimizing metabolic health; no diabetes diagnosis but family history of type 2 diabetes.

#### Artifact-based findings

The system identified a consistent pattern of elevated postprandial glucose excursions (>160 mg/dL) following dinner, temporally associated with late eating (after 9 PM) and poor sleep quality the preceding night. Time-in-range analysis showed 72% overall, but only 58% between 6 PM and midnight. The artifact captured this as a derived insight with source attribution to CGM data, sleep tracker, and meal logs.

#### Generated recommendation

“*CGM data reveals a pattern of evening glucose excursions associated with late meals and sleep quality. Consider: (1) earlier dinner timing (before 7 PM), (2) assessment of sleep-disordered breathing given correlation between poor sleep and next-day glucose control, (3) follow-up CGM review in 4 weeks to assess intervention effect*.”

#### Contrast with RAG

The RAG system retrieved passages about diabetes management and generated generic recommendations about “improving glucose control” without engaging with the specific temporal patterns visible in the patient’s data.

### 5.2 Vignette B: Medication Side Effect Detection

#### Patient

58-year-old female initiated on atorvastatin 20 mg for primary cardiovascular prevention 6 weeks prior.

#### Clinical context

Routine follow-up; patient mentions feeling “more tired than usual.”

#### Artifact-based findings

The system’s change-point detection algorithm identified a statistically significant decrease in daily step count (8,400 → 5,200 average) beginning 8 days after statin initiation. Sleep quality metrics showed increased light sleep percentage and decreased deep sleep. The temporal alignment with medication start date was automatically flagged as a potential statin-associated adverse effect.

#### Generated recommendation

“*Activity and sleep data show change point 8 days after atorvastatin initiation: daily steps decreased 38%, deep sleep decreased 22%. Pattern consistent with possible statin-associated myopathy or fatigue. Recommend: (1) CK level, (2) explicit myalgia assessment, (3) consider trial discontinuation if confirmed, with reassessment of cardiovascular risk-benefit*.”

#### Contrast with RAG

The RAG system retrieved information about statin benefits and generated a recommendation to continue therapy without recognizing the temporal relationship between medication start and symptom onset.

### 5.3 Vignette C: Familial Risk Calibration

#### Patient

35-year-old male with father who had myocardial infarction at age 52; patient requesting cardiovascular risk assessment.

#### Clinical context

Multi-omic data available including polygenic risk score for coronary artery disease, lipid panel, inflammatory markers, and coronary calcium score.

#### Artifact-based findings

The system integrated: (1) 85th percentile polygenic risk score for CAD, (2) LDL-C 142 mg/dL (borderline elevated), (3) hs-CRP 2.8 mg/L (intermediate risk), (4) coronary calcium score of 0 (low short-term risk), (5) family history risk factor. The artifact represented these as separate domain facts with integrated risk scoring.

#### Generated recommendation

“*Integrated risk assessment shows elevated genetic susceptibility (PRS 85th percentile) with intermediate phenotypic risk markers (borderline LDL, elevated hs-CRP) but reassuring coronary calcium score of 0. Family history of premature MI increases concern. Recommend: (1) lifestyle optimization (Mediterranean diet, regular exercise), (2) repeat lipid panel and hs-CRP in 6 months, (3) consider coronary calcium score reassessment in 3–5 years, (4) discuss aspirin and statin therapy if lifestyle measures insufficient*.”

#### Contrast with RAG

The RAG system generated a recommendation focused on standard cholesterol management without integrating the polygenic risk score or calcium score data, missing the opportunity for personalized risk calibration.

## 6 Discussion

### 6.1 Summary of Findings

Our findings challenge a core assumption underlying the deployment of RAG systems in clinical contexts: that retrieval augmentation improves factual grounding. In our evaluation, RAG increased unsupported claims by 8.7-fold compared to baseline, while structured artifacts reduced unsupported claims by 40%. These results suggest that the quality of patient data representation—not the quantity of retrieved context—is the primary determinant of factual reliability in clinical LLM applications.

### 6.2 Comparison with Existing Approaches

Our structured artifact approach differs from existing clinical AI architectures in several ways.

#### FHIR-based systems

While FHIR provides excellent syntactic interoperability, it was designed for data exchange rather than LLM consumption. FHIR resources lack the derived insights, temporal aggregations, and explicit provenance chains that our artifacts provide. Systems that simply serialize FHIR resources into LLM context may face similar challenges to raw-text approaches.

#### OMOP-based analytics

The OMOP Common Data Model [37] supports standardized analytics across institutions but focuses on observational research rather than individual patient summarization. Our approach is complementary: OMOP could serve as a source layer feeding artifact compilation.

#### Traditional RAG architectures

Standard RAG systems [11] retrieve based on embedding similarity, which captures semantic relatedness rather than factual applicability. Our results suggest this distinction is critical in clinical contexts where semantically similar text may describe different patients or time points.

#### Agentic medical AI

Recent work on medical AI agents [22] emphasizes conversational interaction and iterative refinement. Our C3 condition incorporates agentic elements but demonstrates that without high-quality representation, agentic complexity provides limited benefit.

### 6.3 Theoretical Framework: Why Representation Reduces Hallucination

We propose an information-theoretic framework explaining our findings. Consider the generation process as information transmission from source data through representation to generated output.

In the raw-text pathway, source data *S* is encoded into clinical notes *N* with some information loss and noise introduction. The LLM then decodes *N* to generate output *O*. Errors accumulate at each stage:

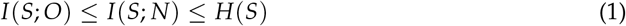

where *I* denotes mutual information and *H* denotes entropy. RAG retrieves a subset *N*_*k*_ ⊂ *N*, potentially reducing *I*(*S*; *N*_*k*_) if retrieval is imperfect.

In the artifact pathway, source data *S* is compiled into structured artifacts *A* designed to preserve clinically relevant information with explicit provenance. The compilation process can actually *increase* accessible information by computing derived features:

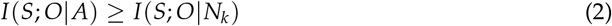

when the artifact compilation process extracts signal from noise and makes implicit relationships explicit.

Furthermore, the structured format reduces the *decoding complexity* for the LLM. Natural language clinical notes require the model to perform implicit entity resolution, temporal reasoning, and uncertainty interpretation. Structured artifacts make these explicit, reducing the computational burden and error opportunity during generation.

This framework predicts that RAG failures should be more common when: (1) semantic similarity diverges from factual relevance, (2) source documents have high lexical overlap despite describing different entities, and (3) temporal or contextual information is implicit in the text. Clinical documentation exhibits all three characteristics, explaining the poor RAG performance in our evaluation.

### 6.4 Clinical Implications

Our findings have implications for healthcare systems deploying clinical AI:

#### Investment priorities

Resources directed toward improving data representation may yield greater safety returns than optimizing retrieval systems or adding workflow complexity. Healthcare organizations should consider investing in data infrastructure that produces high-quality structured patient representations.

#### Regulatory frameworks

FDA guidance on AI/ML-based medical devices [45] emphasizes transparency and validation. Structured artifacts with explicit provenance support these goals by enabling automated consistency checking and auditability.

#### Clinical workflow integration

The artifact-based approach supports local-first deployment (14% smaller context) with better performance, potentially enabling privacy-preserving architectures where patient data need not leave institutional boundaries.

#### Human-AI collaboration

The agent workflow’s escalation mechanism (12% of cases in C3) provides a principled approach to human-in-the-loop verification for uncertain or high-stakes cases, though calibration of escalation thresholds requires further study.

### 6.5 Ethical Considerations

The deployment of AI systems for clinical decision support raises important ethical considerations:

#### Data privacy

Multi-source health data integration, including wearables and genomics, requires robust consent frameworks and data governance. The local-first potential of artifact-based systems may help address privacy concerns by reducing data transmission.

#### Algorithmic bias

Artifact compilation processes may encode biases present in source systems (*e*.*g*., differential documentation quality across demographic groups). Evaluation across diverse populations is essential before deployment.

#### Informed consent

Patients should understand when AI systems contribute to their care and have opportunities to opt out or request human-only review.

#### Professional responsibility

AI-assisted recommendations must ultimately be validated by qualified clinicians. System design should support rather than replace clinical judgment.

#### Equity of access

Advanced AI decision support should not exacerbate existing healthcare disparities. Deployment strategies should consider access across different care settings and populations.

### 6.6 Limitations

Several limitations constrain interpretation of our findings:

#### Synthetic data

All vignettes were synthetically generated rather than derived from real patient records. While designed to reflect realistic complexity, synthetic data may not capture the full heterogeneity of real-world clinical documentation. Validation on real EHR data with appropriate privacy protections is essential.

#### Single model architecture

All experiments used GPT-4o-mini. Results may differ with other architectures, model sizes, or domain-adapted models. Larger models might handle retrieval integration more robustly; specialized medical models might show different patterns.

#### Gold standard derivation

Gold-standard facts derived from structured artifacts potentially bias evaluation toward artifact conditions. We mitigated this by having clinicians verify gold standards independently of artifact representation.

#### Limited scale

100 vignettes across 3 seeds may miss rare failure modes. Larger-scale evaluation would strengthen confidence in findings.

#### Single RAG configuration

We evaluated one RAG implementation with *k* = 10 retrieval. Alternative configurations (different *k*, reranking, hybrid retrieval) might yield different results, though our analysis suggests the fundamental semantic-vs-factual relevance problem would persist.

#### Evaluation by automated system

Primary outcome measurement used GPT-4o-mini for claim extraction and verification. While we validated inter-rater reliability with human review, fully automated evaluation may miss subtle errors.

### 6.7 Future Directions

Several directions warrant further investigation:

#### Real-world validation

Evaluation on de-identified real clinical data would strengthen confidence and reveal failure modes specific to production healthcare environments.

#### Multi-model evaluation

Comparing findings across model families (GPT-4, Claude, Gemini, open-source medical models) would establish generalizability.

#### Hybrid approaches

Combining structured artifacts with selective retrieval for specific query types might capture benefits of both approaches.

#### Prospective clinical studies

Evaluating impact on clinical workflows, physician efficiency, and ultimately patient outcomes would demonstrate clinical utility.

#### Artifact compilation optimization

Research into optimal artifact schemas, update frequencies, and domain ontologies could further improve representation quality.

## 7 Conclusion

RAG increased hallucination nearly 9-fold in clinical text generation, contradicting assumptions underlying its widespread adoption in healthcare AI. Structured patient artifacts with explicit provenance substantially reduced unsupported claims while enabling smaller context footprints suitable for privacy-preserving deployment.

Our findings suggest that representation quality, not retrieval quantity, determines the reliability of clinically-grounded language model outputs. The information-theoretic advantage of structured representation—making implicit clinical relationships explicit while preserving provenance—explains this advantage.

Development of trustworthy clinical AI should prioritize structured data representation over retrieval augmentation. Investment in patient-state compilation infrastructure may yield greater safety returns than optimization of retrieval systems or elaboration of agentic workflows. As healthcare systems increasingly integrate AI into clinical workflows, the foundation of reliable structured representation becomes essential for achieving the promise of AI-assisted medicine while maintaining the safety standards patients deserve.

## Author Contributions

**JS** conceived the system architecture, designed the experimental framework, conducted the experiments, performed statistical analyses, and drafted the manuscript. **AC** provided clinical expertise, developed the evaluation framework for clinical safety and utility, conducted physician reviews, and contributed to manuscript revision. **MV** provided strategic direction, healthcare systems perspective, and critical revision of the manuscript. All authors approved the final version.

## Data Availability

All data produced in the present study are synthetic. Synthetic vignette specifications, artifact schema, and evaluation code will be made available in a public repository upon publication. No real patient data was used in this study.

https://github.com/joescanlin/myome-cds

## Data Availability

Synthetic vignette specifications and evaluation code will be made available in a public repository upon publication. The artifact schema specification is provided in Appendix A. Real patient data was not used in this study.

## Acknowledgments

We thank the clinical advisors who reviewed vignette realism and clinical scenario validity. We thank the board-certified physicians who conducted blinded safety reviews. We thank early readers for feedback that improved the manuscript.

## A Artifact Schema Specification

### A.1 Core Data Structures

**Listing 1:**
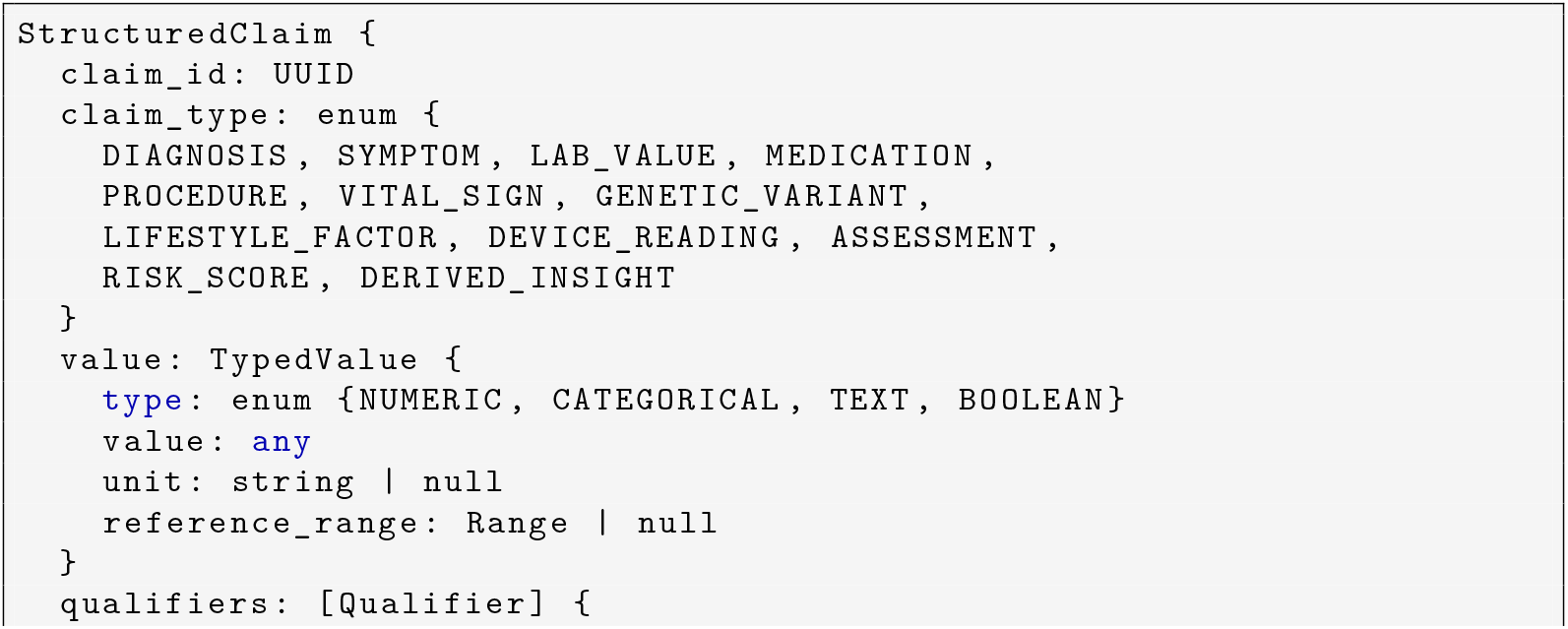
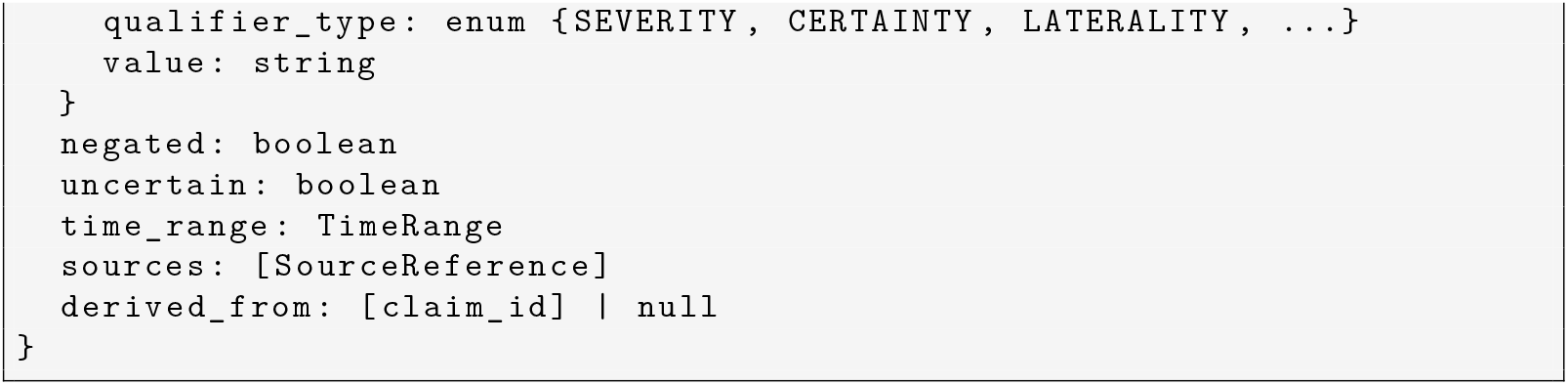
StructuredClaim Schema

**Listing 2:**
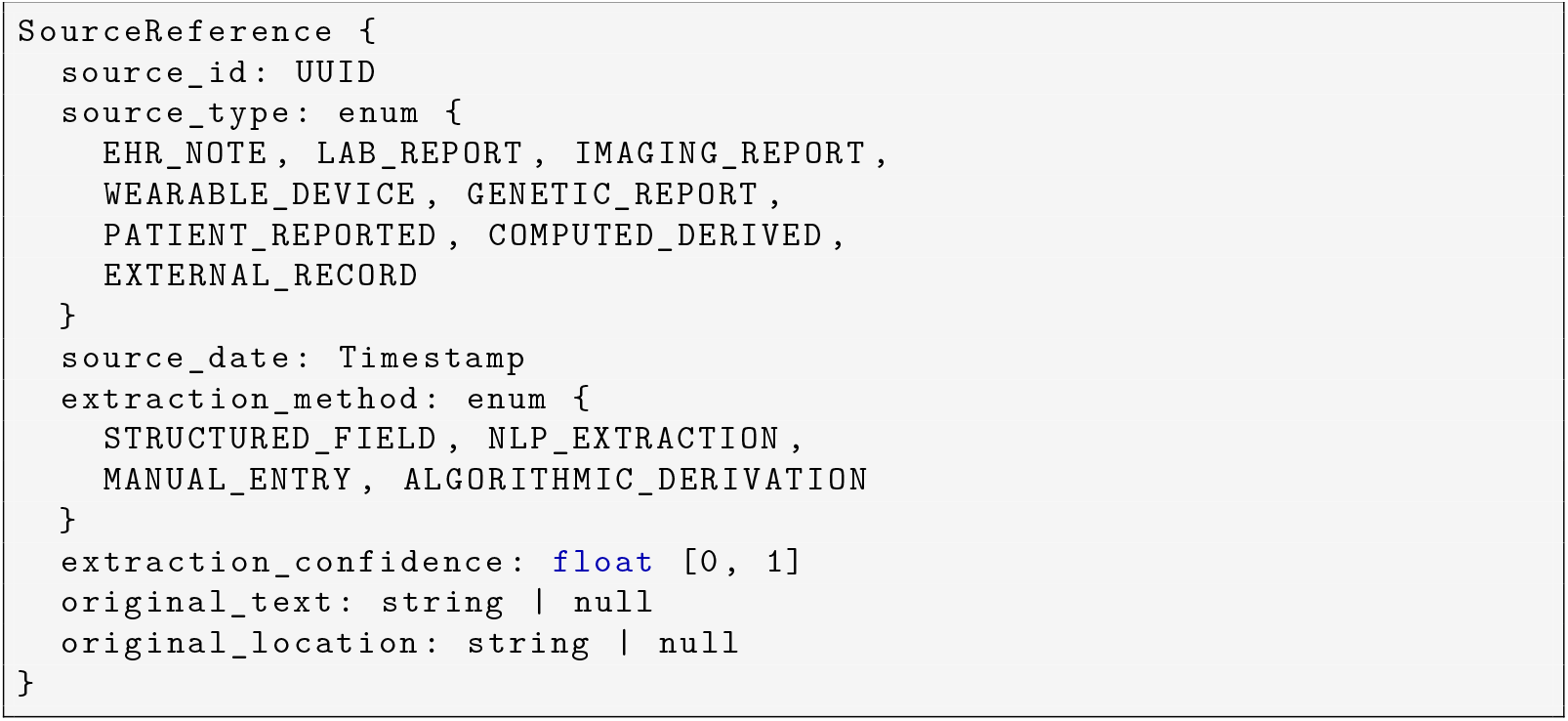
SourceReference Schema

**Listing 3:**
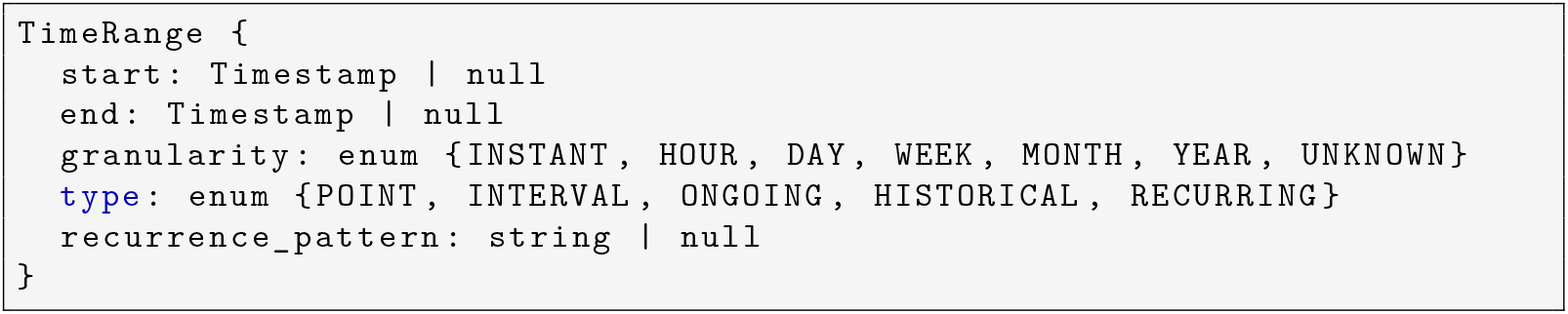
TimeRange Schema

### A.2 Validation Rules

Artifacts undergo validation before use in generation:

1. **Referential integrity:** All `derived_from` references resolve to existing claims within the artifact.
2. **Temporal consistency:** No claim references a source dated after the claim’s valid time end.
3. **Ontological validity:** All coded values (diagnoses, medications, procedures) map to recognized terminologies (ICD-10, RxNorm, CPT, LOINC).
4. **Completeness:** Required fields present for each claim type as defined by domain-specific schemas.
5. **Confidence bounds:** Extraction confidence scores within [0, 1] and consistent with extraction method.

## B Clinical Vignette Construction

### B.1 Synthetic Vignette Generation Process

Synthetic vignettes were generated through a structured process:

1. **Scenario selection:** Clinical scenarios sampled from predefined distributions across categories (chronic disease, acute presentation, preventive care, multi-morbidity).
2. **Demographic generation:** Patient demographics (age, sex, relevant social history) generated with realistic distributions.
3. **Condition assignment:** Primary and comorbid conditions assigned based on epidemiological plausibility and scenario requirements.
4. **Data source simulation:** For each condition and data type, synthetic records generated including: EHR notes, lab values, vital signs, wearable data streams, and medication lists.
5. **Complexity injection:** Realistic complexity factors added: documentation gaps, conflicting information between sources, borderline findings, temporal evolution.
6. **Gold standard creation:** Clinician-verified fact sets created for each vignette, reviewed independently by two physicians with disagreements resolved by discussion.

### B.2 Example Vignette: Chronic Disease Management

**Scenario:** 58-year-old female with type 2 diabetes, hypertension, and chronic kidney disease stage 3.

#### Complexity factors

- Conflicting blood pressure readings between home monitoring and clinic visits
- Recent medication change with incomplete documentation
- Wearable data showing nocturnal hypoglycemia not captured in HbA1c

#### Gold standard facts (selected)

- Type 2 diabetes diagnosed 2019
- Current HbA1c 7.8% (measured 2024-01-15)
- eGFR 42 mL/min/1.73m^2^ (measured 2024-01-10)
- Metformin 1000 mg twice daily (current)
- Lisinopril 20 mg daily discontinued 2023-11-20 due to hyperkalemia
- Home systolic BP range 128–156 mmHg over past month
- CGM data shows 3.2 hypoglycemic episodes per week (glucose *<*70 mg/dL) between 2–5 AM

## C Supplementary Results

### C.1 Results by Vignette Category

### C.2 Inter-Rater Reliability Details

For physician safety review:

- Raw agreement: 94.2%
- Cohen’s *κ*: 0.78 (95% CI: 0.64–0.92)
- Disagreements (*n* = 14): Resolved by consensus discussion
- Primary disagreement sources: Severity assessment of documentation gaps, clinical significance of minor temporal errors

For claim extraction verification (10% sample):

- Raw agreement: 91.8%
- Cohen’s *κ*: 0.82 (95% CI: 0.71–0.91)

### C.3 Confidence Calibration Analysis

Model-expressed confidence correlated poorly with accuracy in RAG: high-confidence claims were incorrect 38% of the time. Artifact conditions showed better calibration, with high-confidence errors at 7% (C2) and 14% (C3).

